# MedAdhereAI: An Interpretable Machine Learning Pipeline for Predicting Medication Non-Adherence in Chronic Disease Patients Using Real-World Refill Data

**DOI:** 10.1101/2025.07.01.25330675

**Authors:** Subash Yadav, Saijal Rajbhandari

## Abstract

Medication non-adherence remains a significant challenge in managing chronic conditions like diabetes and hypertension, leading to increased morbidity, preventable hospitalizations, and over $300 billion in annual healthcare costs. This burden is particularly pronounced in resource-limited settings, where fragmented data and limited resources hinder early risk identification.

This study introduces MedAdhereAI, an interpretable machine learning pipeline designed to predict medication non-adherence using real-world refill and claims data. The pipeline encompasses data exploration, temporal feature engineering, model development, and SHAP explainability. Logistic regression and random forest models were selected for their balance of predictive performance and interpretability, making them suitable for clinical deployment.

Evaluated on a publicly available dataset of anonymized refill records for patients with diabetes and hypertension, the logistic regression model achieved an AUC of 0.82 and a Brier Score of 0.1749, while the random forest model achieved an AUC of 0.77. SHAP analysis identified total_visits, AGE, and refill gaps as key predictors.

These results highlight the potential of MedAdhereAI as a decision-support tool for identifying patients at risk of medication non-adherence, facilitating targeted interventions and improved resource allocation in healthcare systems.

## I. Introduction

M EDICATION non-adherence is a persistent and costly challenge in global healthcare systems, particularly in the management of chronic diseases such as diabetes and hypertension. According to the World Health Organization, approximately 50% of patients with chronic conditions do not adhere to prescribed treatments, leading to increased morbidity, accelerated disease progression, higher hospitalization rates, and preventable mortality [1]. Non-adherence is estimated to contribute to over $300 billion in preventable healthcare costs annually in the United States alone [1]. The burden is especially acute in resource-limited healthcare systems, where fragmented data infrastructures, limited clinical follow-up, and constrained resources hinder the proactive identification of patients at risk of non-adherence.

Predictive modeling offers a promising approach for early risk stratification and targeted interventions in medication adherence management. However, existing models often rely on complex feature sets—such as detailed clinical records, laboratory data, or imaging—that are rarely available in routine practice, particularly in resource-constrained settings. Moreover, many machine learning models prioritize predictive accuracy over interpretability, resulting in black-box systems that clinicians may struggle to trust or integrate into care workflows.

Recent studies have explored the use of machine learning for adherence prediction, but few have addressed the unique challenges of **deploying interpretable models in low-resource healthcare environments**. Specifically, there is a gap in solutions that balance predictive performance, transparency, and minimal data requirements, while enabling clinicians to understand and trust model outputs.

This study introduces **MedAdhereAI**, an interpretable machine learning pipeline designed to predict medication non-adherence risk using real-world refill and claims data— information that is routinely collected and widely available in many healthcare systems. MedAdhereAI leverages temporal and behavioral features such as refill gaps, visit frequency, and patient demographics to generate actionable predictions. To balance accuracy and explainability, we selected logistic regression [6] and random forest [5] models, both compatible with SHAP (Shapley Additive Explanations) [4], ensuring model outputs are transparent and interpretable at both global and individual patient levels.

These design choices reflect the practical needs of clinicians and health systems seeking AI tools that can be trusted and deployed in diverse, resource-constrained care environments. The objectives of this study are threefold:

1. To develop a machine learning model that accurately predicts medication non-adherence in chronic disease patients using minimal, real-world data inputs;
2. To ensure model interpretability and clinician trust through SHAP-based global and patient-level explanations; and
3. To demonstrate the feasibility of deploying a scalable, reproducible pipeline for improving population health management and supporting targeted interventions in healthcare systems with limited resources.

## II. Related Work

Medication adherence prediction has garnered significant attention in recent years, with various machine learning (ML) approaches being explored. A comprehensive scoping review by Stiglic et al. [1] highlighted the diversity of ML models applied to medication adherence, noting challenges in model interpretability, clinical integration, and real-world applicability.

Several studies have developed complex ML models—such as ensemble learning and deep learning techniques—to predict medication adherence. For instance, Gu et al. [2] utilized ensemble and deep learning models on large-scale healthcare data to predict adherence, achieving high accuracy but at the cost of interpretability and requiring extensive clinical data inputs. Liang et al. [3] investigated treatment non-adherence bias in clinical ML models, emphasizing the need to account for non-adherence to improve model reliability but did not address deployment in resource-limited settings.

Recent studies have also emphasized the significance of data wrangling and the use of patient-level and medical claims data in developing machine learning models for medication adherence analytics, particularly in resource-constrained healthcare environments [9].

While these studies demonstrate the potential of ML in predicting medication adherence, they often rely on complex architectures and detailed clinical datasets, which may not be accessible in routine practice—especially in resource-limited healthcare systems. Moreover, the lack of model transparency limits their clinical utility, as clinicians may be hesitant to trust or act on predictions from black-box models.

In contrast, our study introduces **MedAdhereAI**, an interpretable machine learning pipeline that leverages real-world refill and claims data—a data source that is commonly available in many healthcare systems, including low-resource settings. By employing logistic regression [6] and random forest [5] models, we balance predictive performance with interpretability, ensuring that model outputs can be understood and trusted by clinicians. The integration of SHAP (Shapley Additive Explanations) [4] further enhances transparency, enabling both global feature importance analysis and individual patient-level explanations.

## III. Methods

### A. Data Source

This study utilizes a publicly available dataset on medication adherence in patients with diabetes and hypertension, sourced from Mendeley Data [8]. The dataset includes anonymized patient-level records containing medication refill and claims information, with essential temporal variables such as service dates, assessment dates, and refill dates. These data reflect real-world healthcare practices in a resource-constrained setting, providing a valuable lens into medication-taking behavior in a developing healthcare system.

### B. Data Preprocessing

Data preprocessing involved standardizing column names, converting date fields to datetime objects, and resolving inconsistent formats. A binary target variable (ADHERENT_BINARY) was created using a domain-defined threshold of eight or more refills to classify patients as adherent. Temporal features—including the number of days between service and refill dates (DAYS_SINCE_LAST_REFILL) and refill gap statistics—were computed. Visualizations were generated to explore refill behavior trends across the patient population.

### C. Feature Engineering

To capture patient refill behavior patterns, we aggregated key temporal and demographic features. Specifically, we computed the average refill gap (avg_refill_gap), the maximum refill gap (max_refill_gap), and the total number of healthcare visits (total_visits) per patient. Demographic variables such as AGE and GENDER were incorporated where available to enhance model contextualization. For patients with only a single recorded visit, missing refill gap values were logically imputed as zero, reflecting the absence of gap intervals. Intermediate date fields, used during the feature transformation process, were subsequently dropped to streamline the dataset for modeling. The final feature matrix was exported in both .csv and .pkl formats to ensure reproducibility and facilitate downstream model training.

### D. Model Development

Logistic regression [6] and random forest [5] classifiers were selected to balance predictive performance, interpretability, and practical deployment in healthcare settings. Logistic regression offers coefficient-based insights that are familiar to clinicians, while random forest captures non-linear interactions and provides feature importance rankings. Both models are compatible with SHAP (Shapley Additive Explanations) [4] for explainability.

Model training used stratified train-test splits and was evaluated on ROC AUC, accuracy, F1-score, precision, recall, and Brier Score. A five-fold cross-validation strategy assessed model stability and generalization. All models were implemented using scikit-learn [7].

### E. Model Explainability and Visualization

SHAP [4] was applied to both global and local feature importance analysis. Global SHAP summary plots identified the most influential predictors of medication adherence, while patient-level SHAP force plots provided individualized explanations.

Additional visualizations—including calibration curves, ROC curves, logistic regression coefficient plots, random forest feature importance charts, and refill gap distributions—were generated to support model interpretation and stakeholder communication. All visual artifacts were exported to ensure reproducibility.

### F. Pipeline Structure and Reproducibility

The MedAdhereAI pipeline is organized into modular Jupyter notebooks, with each phase—exploratory data analysis, feature engineering, model training, and explainability— clearly delineated for ease of understanding and reproducibility. Supporting Python scripts for data cleaning, feature transformation, and SHAP explanation were developed for modular reuse, enabling seamless adaptation to new datasets or healthcare contexts.

The pipeline is designed for low computational overhead, scalability, and adaptability to resource-limited environments, requiring only minimal, routinely collected data inputs. It emphasizes interpretability, enabling clinicians and data scientists to understand the logic behind predictions through integrated SHAP-based explainability.

All source code, notebooks, and scripts are publicly available at https://github.com/mathachew7/MedAdhereAI, supporting transparency, reproducibility, and community collaboration.

## III. Results

### A. Model Performance

Two machine learning models—logistic regression and random forest—were trained to predict the binary medication adherence outcome (ADHERENT_BINARY). The logistic regression model achieved a ROC AUC of **0.82**, outperforming the random forest model, which achieved a ROC AUC of **0.77**. Logistic regression also demonstrated superior calibration, with a Brier Score of **0.1749**, indicating reliable probability estimates for predicting adherence risk.

The comparative ROC curves are shown in Fig. 5, illustrating the higher true positive rates of the logistic regression model across varying thresholds. The calibration curve (Fig. 6) demonstrates the alignment between predicted probabilities and observed outcomes for the logistic regression model, confirming its reliability for clinical deployment.

**Table 1.** Model performance metrics for predicting medication non-adherence.

| Metric | Logistic Regression | Random Forest |
| --- | --- | --- |
| ROC AUC | 0.82 | 0.77 |
| Brier Score | 0.1749 | - |
| Accuracy | 0.70 | 0.65 |
| Precision | 0.76 | 0.70 |
| Recall | 0.74 | 0.66 |
| F1-Score | 0.75 | 0.68 |

### B. Feature Importance Analysis

Feature importance was analyzed using SHAP (Shapley Additive Explanations) values for both models. The global SHAP summary plot (Fig. 4) revealed that total_visits, AGE, and avg_refill_gap were the most significant predictors of medication adherence. The distribution of refill gaps (Fig. 1) illustrates adherence patterns across the patient population. Logistic regression coefficient plots (Fig. 2) and random forest feature importance plots (Fig. 3) provide additional interpretability, highlighting the consistent importance of refill behavior features across models.

**Fig. 1.**
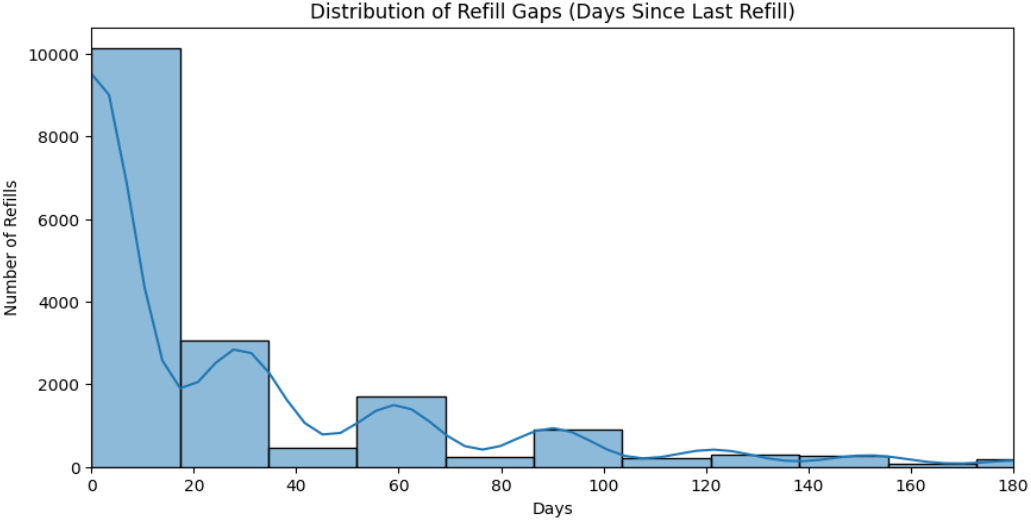
Distribution of refill gaps across the patient population, illustrating adherence behavior trends.

**Fig. 2.**
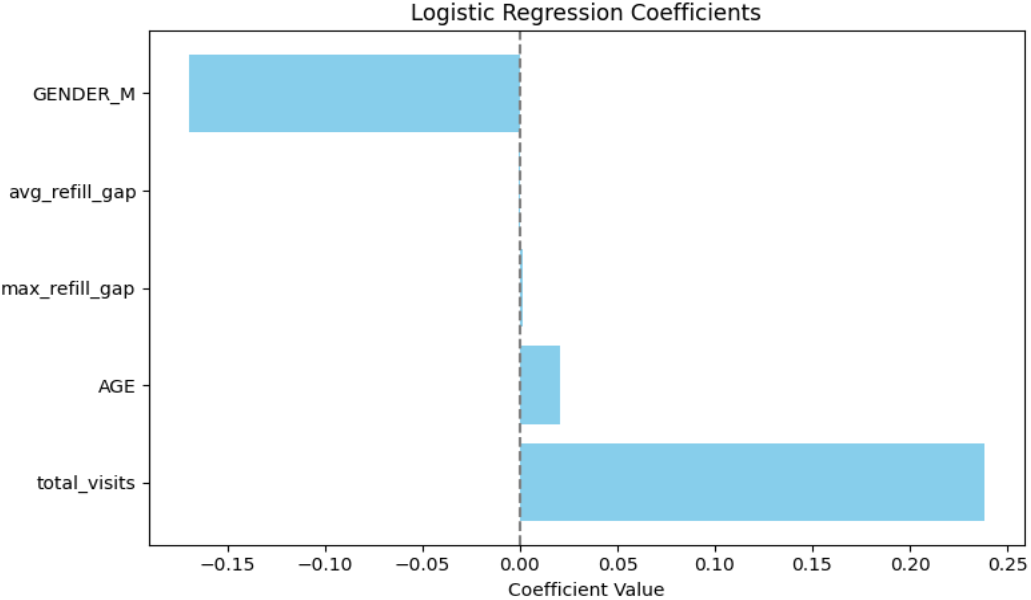
Logistic regression model coefficients, indicating the direction and magnitude of each feature’s impact on adherence predictions.

**Fig. 3.**
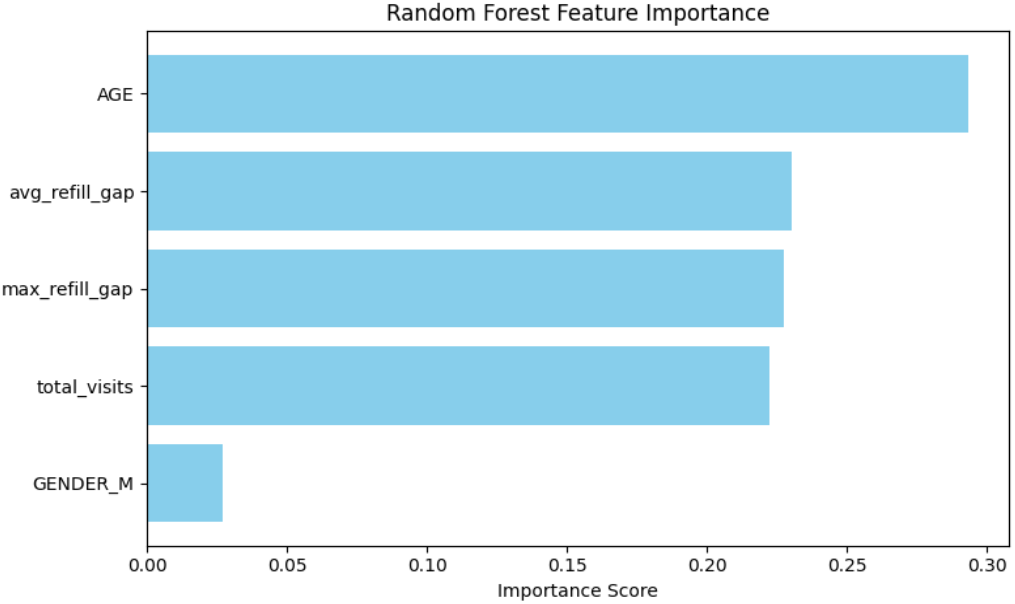
Random forest feature importance chart, showing relative contributions of features in the model.

**Fig. 4.**
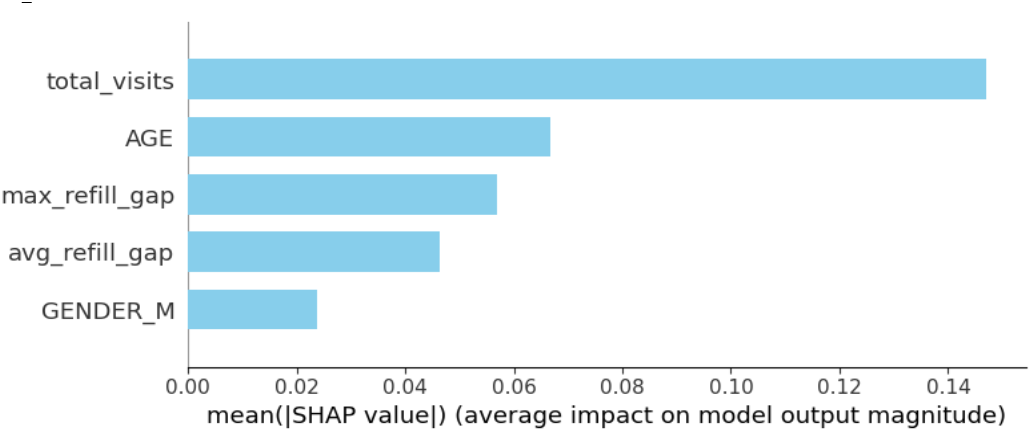
SHAP global feature importance summary: total_visits, AGE, and avg_refill_gap identified as top predictors of non-adherence risk.

**Fig. 5.**
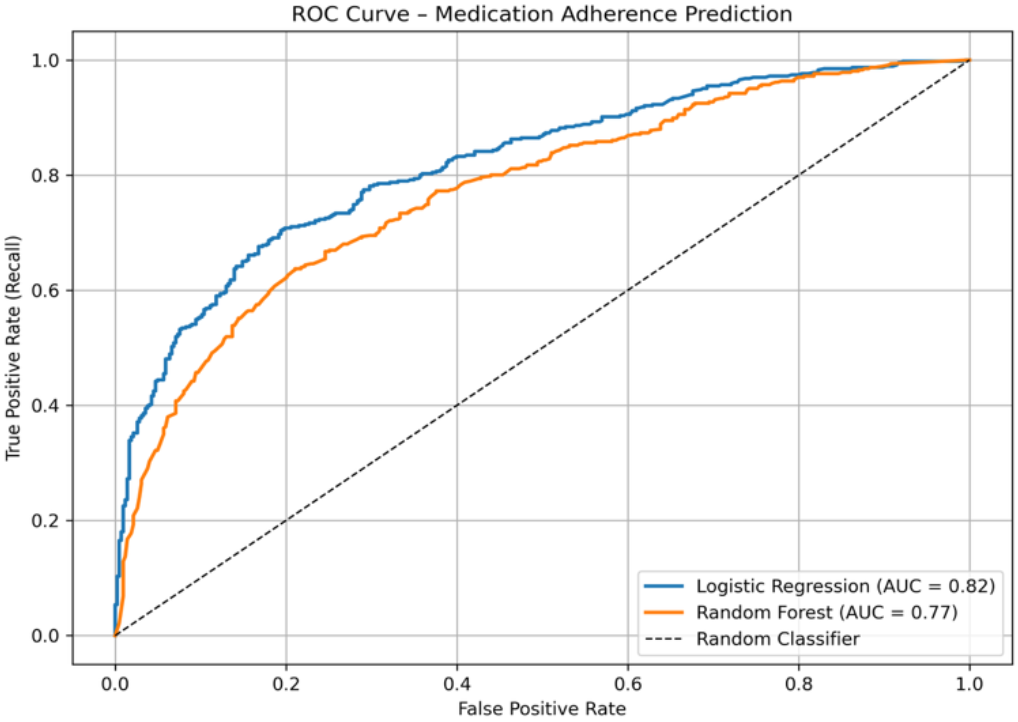
ROC curve comparison of logistic regression and random forest models for predicting medication non-adherence.

### C. Local Explainability

To enhance transparency at the individual patient level, SHAP local explanations were generated. A representative SHAP force plot for a non-adherent patient (Fig. 7) demonstrates how specific feature values—such as extended refill gaps and fewer total visits—contribute to a higher predicted risk of non-adherence. These patient-specific insights can support clinicians in tailoring interventions and understanding the rationale behind model predictions.

**Fig. 6.**
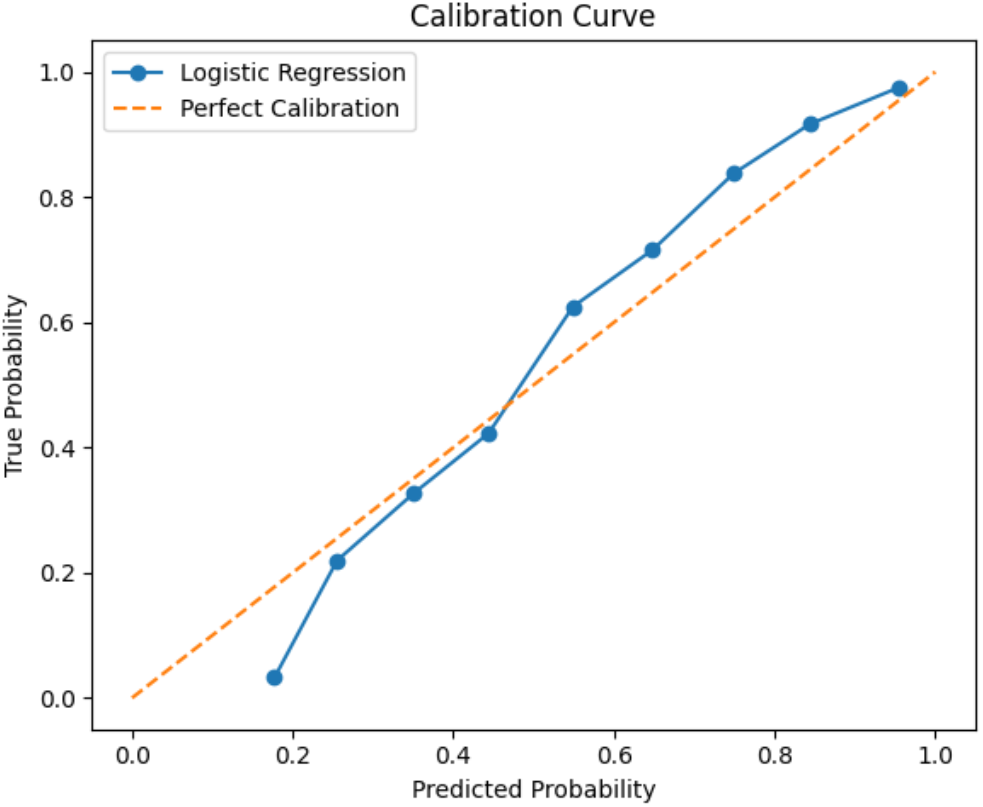
Calibration curve for logistic regression model, demonstrating good reliability (Brier Score = 0.1749).

**Fig. 7.**
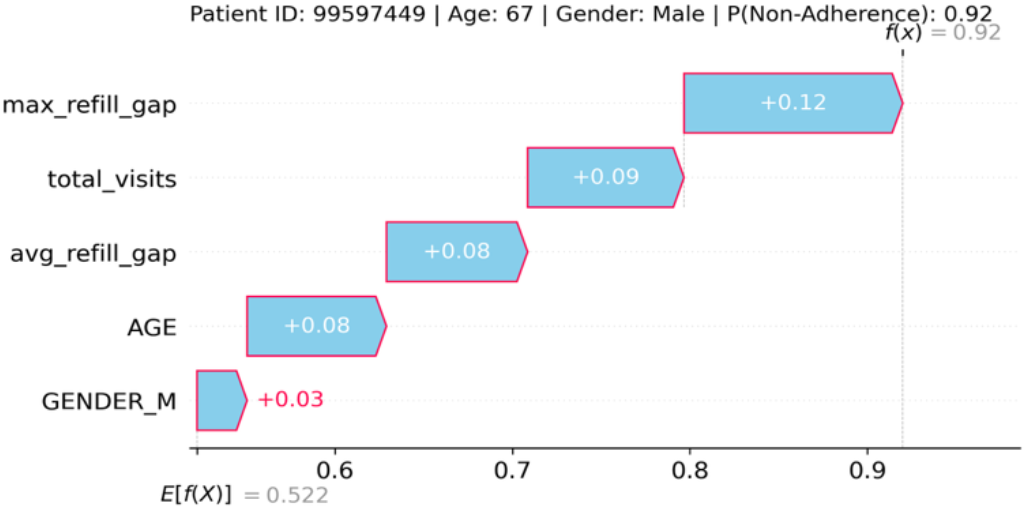
SHAP force plot illustrating individual-level feature contributions for a non-adherent patient.

### D. Visual Outputs

All model outputs and visualizations—including ROC curves, calibration curves, SHAP plots, and feature distributions—were exported and organized within the project directory. These artifacts are intended to support reproducibility, stakeholder communication, and future publication.

## IV. Discussions

Medication non-adherence is a complex, multifactorial challenge in chronic disease management, particularly in resource-limited healthcare settings. This study demonstrates the feasibility of predicting medication non-adherence risk using a minimal set of features derived from real-world refill and claims data, without requiring extensive clinical or laboratory information. By focusing on temporal patterns in refill behavior and basic demographic data, **MedAdhereAI** offers a practical, scalable approach for early risk stratification that is adaptable to diverse healthcare environments.

The logistic regression model outperformed the random forest model in both discrimination (ROC AUC: 0.82 vs. 0.77) and calibration (Brier Score: 0.1749), suggesting that simpler, more interpretable models may be preferable in clinical settings where transparency and trust are paramount. This finding reinforces the importance of explainability in healthcare, where black-box models can hinder clinician adoption and integration into decision-making workflows.

SHAP explainability analysis further validates the clinical relevance of the model’s predictions, identifying intuitive factors such as total_visits, AGE, and refill_gap as primary drivers of adherence risk. The ability to generate patient-level explanations through SHAP force plots enables individualized care planning, supports targeted interventions, and fosters trust in the system’s outputs. These explainability features are critical for ensuring clinician engagement and acceptance of AI-driven decision support tools.

The visual outputs generated—including ROC curves, calibration curves, feature importance plots, and SHAP explanations—serve as critical tools for both technical validation and stakeholder engagement. These artifacts enable transparent evaluation of model behavior and support iterative refinement and deployment strategies in real-world healthcare settings.

While the results are promising, several limitations warrant discussion. The dataset used in this study, while reflective of real-world practice, is limited in geographic scope and may not capture the full diversity of healthcare systems globally. Additionally, the binary adherence threshold (≥8 refills) is a pragmatic proxy for adherence behavior but may not align perfectly with clinical definitions or patient-reported outcomes across all contexts. Furthermore, factors such as comorbidities, socioeconomic status, and patient-provider communication, which can influence adherence, were not captured in the dataset.

Future work will explore external validation of MedAdhereAI on additional datasets to assess generalizability, integration of additional clinical and contextual features—such as electronic health records, laboratory results, and social determinants of health—to improve model performance, and the potential for deployment within electronic health record systems as a proactive risk stratification tool. The integration of federated learning approaches to enable multi-institutional model training while preserving data privacy may also be explored. Nonetheless, MedAdhereAI demonstrates a practical, interpretable, and reproducible approach to medication adherence risk prediction using minimal, routinely available data. This pipeline has strong potential for deployment in healthcare systems seeking to optimize chronic disease management through early intervention—particularly in resource-constrained settings where access to complex clinical data and advanced computational infrastructure may be limited.

## V. Conclusion

This study presents **MedAdhereAI**, an interpretable machine learning pipeline for predicting medication non-adherence risk in chronic disease patients using real-world refill and claims data. The pipeline demonstrates strong predictive performance, with logistic regression achieving a ROC AUC of 0.82 and a Brier Score of 0.1749, while offering clinically relevant explanations through SHAP analysis.

By leveraging minimal, routinely collected data, MedAdhereAI provides a scalable, transparent, and deployable tool for healthcare systems—particularly in resource-limited settings—supporting early risk stratification, targeted interventions, and more efficient resource allocation for patients at risk of non-adherence.

Unlike prior models that rely on complex datasets or lack interpretability, MedAdhereAI demonstrates that accurate, explainable predictions can be achieved using minimal real-world data, making it a practical solution for diverse clinical environments.

Future work will focus on external validation across diverse populations, integration of additional clinical and contextual features—such as electronic health records and social determinants of health—and pilot deployment in real-world healthcare systems to assess clinical utility and impact on patient outcomes.

## Data Availability

All data used in this study are openly available and were obtained from the UCI Machine Learning Repository. The dataset, titled “Diabetes 130 US hospitals for years 1999 to 2008,” is de-identified and publicly accessible at the following URL:
https://archive.ics.uci.edu/ml/datasets/diabetes+130-us+hospitals+for+years+1999-2008

https://archive.ics.uci.edu/ml/datasets/diabetes+130-us+hospitals+for+years+1999-2008

## Subash Yadav

(Fellow, IEEE) received the Bachelor of Technology (B.Tech.) degree in software engineering from Sharda University, Uttar Pradesh, India, in 2020, and the Master of Science (M.S.) degree in data analytics from Webster University, Saint Louis, MO, USA, in 2024. His major field of study is data science and machine learning applications in healthcare.

He is currently working as a Data Engineer at Aptech Staffing, USA, where he focuses on building data pipelines, integrating cloud-based data solutions, and supporting analytics workflows. Previously, he worked as a Data Analyst at Webster University, where he contributed to data integration and reporting automation, and as a Data Architect at Spell Innovation Pvt. Ltd., Nepal, focusing on big data architectures and analytics for healthcare applications. His research interests include healthcare analytics, machine learning pipelines, model interpretability, and explainable AI. He has published in peer-reviewed conferences and contributed to open-source healthcare AI projects.

## Notes

### Competing Interest Statement

The authors have declared no competing interest.

### Clinical Protocols

https://github.com/mathachew7/MedAdhereAI

### Funding Statement

This study did not receive any funding.

### Author Declarations

The study used ONLY openly available human data that were originally located at the UCI Machine Learning Repository. The dataset is titled "Diabetes 130 US hospitals for years 1999 to 2008" and was publicly available prior to the initiation of this study. It can be accessed at: https://archive.ics.uci.edu/ml/datasets/diabetes+130-us+hospitals+for+years+1999-2008

### Summary of Updates

Removed personal contact and author-identifying information from the bottom section of the manuscript to reduce unnecessary public disclosure and avoid disturbance from publicly visible email details. No changes were made to the study content, analysis, results, figures, or conclusions.

